# Remifentanil as an alternative in modified rapid sequence induction: A Proof-of-concept study in a selected paediatric population

**DOI:** 10.1101/2024.12.11.24318829

**Authors:** Julien Pico, Chrystelle Sola, Laurent Hertz, Julien Riera, Christopher Scott, Kévin Chapron, Philippe Pirat, Sophie Bringuier, Christophe Dadure

## Abstract

**Introduction:** Rapid sequence induction (RSI) is widely used in paediatric anaesthesia. Concerns over complications associated with classical RSI have prompted the exploration of alternatives. This study was conducted to determine the feasibility and safety of using bolus remifentanil in combination with a hypnotic agent for paediatric RSI.

**Methods:** This proof-of-concept study analysed data from paediatric patients, with at least one risk factor for pulmonary aspiration, undergoing RSI with remifentanil at the Montpellier University Hospital between December 2021 and August 2023. Exclusion criteria included the planned use of a neuromuscular blocking agent for RSI, preoperative hypoxemia, hemodynamic shock or difficult intubation risk factors. Remifentanil was administered by direct intravenous injection with optional prophylactic administration of atropine. Data on intubation success rates, major and minor complications and rescue treatment were collected and analysed.

**Results:** The study included 267 children with a mean age of 7.8 +/- 4.4 years. The success rate for the first intubation attempt was 92.9% (95% CI, 88.9-95.6). No major complication or pulmonary aspiration was reported. Minor complications, primarily hypotension, occurred in 15.7% of patients. The prophylactic use of atropine was correlated with a reduction in minor complications.

**Discussion:** This study supports the use of remifentanil for RSI in a selected paediatric population. Remifentanil offers good intubation conditions with a high success rate at the first attempt and a high safety profile with no major complication reported. Prophylactic atropine administration seems beneficial in reducing minor complications. These findings advocate for integrating remifentanil as an alternative in protocols for modified RSI. Further randomised studies are required to validate such outcomes and refine clinical approaches.

## INTRODUCTION

As pulmonary aspiration remains a potentially devastating complication of general anaesthesia (GA) in all age groups (1), rapid sequence induction (RSI) is a standard procedure for tracheal intubation in children at risk of aspiration or regurgitation of gastric contents. Current data from the APRICOT study (2) show that almost 11% of paediatric general anaesthesia are performed by RSI, making this a regular practice of paediatric anaesthesiologists.

As in adults, the aim of RSI in the paediatric population is to achieve excellent intubation conditions and to minimise the delay between loss of consciousness and airway protection by tracheal intubation. Reduced apnoea tolerance combined with reduced functional residual capacity (3) in children lead to an increased risk of hypoxemia: the younger the child, the greater the risk of desaturation.

The French guidelines, entitled “*Management of the child’s airway under anaesthesia”* (4), recommend using a rapid intravenous neuromuscular blocking agent during a conventional RSI (Grade 1+), with succinylcholine being the first choice. Experts introduce the concept of a “modified” or “controlled” RSI with a gentle mask ventilation not exceeding 15 cm H2O to maximise oxygen reserves and to reduce the risk of hypoxemia before intubation.

In current practice, according to Weiss et al. (5), the prevention of pulmonary aspiration, which remain rare, even in paediatrics, is wrongly considered more important than limiting the risk of hypoxemia and other related complications (6). Many authors now consider that there is a wide range of possible practices for paediatric RSI. The current debate between succinylcholine and rocuronium seems to be obsolete, as the ‘‘classic” RSI should be abandoned in paediatric anaesthesia for its ‘‘controlled’’ approach (7–10). In contrast to serious respiratory events, which are still frequent in paediatric anaesthesia (1), the incidence of pulmonary aspiration is low, observed in 1 in 4498 cases in a prospective study conducted over one year in specialised paediatric centres (11).

Remifentanil, administered as an intravenous (IV) bolus, is an ultra-short-acting opioid with interesting pharmacokinetic properties. The elimination is extremely rapid, through tissue and plasma esterases. The volume of distribution is increased in children, especially in neonates, and clearance is even faster, unlike other opioids (12).

Remifentanil used as an IV injection of 3 to 4µg/kg appears to offer optimal intubation conditions based on a scoring system (13) in a paediatric population without causing severe cardiovascular depression or signs of muscular rigidity (14).

The aim of this study was to evaluate the feasibility and safety of using of IV bolus remifentanil in combination with a hypnotic agent in a selected paediatric population undergoing RSI.

## METHODS

### Study design

We conducted a retrospective analysis of prospectively recorded data, in the department of paediatric anaesthesia of the Montpellier University Hospital. Anonymised anaesthesia data from children who received remifentanil for RSI between December 2021 and August 2023 were collected.

### Population

Children aged up to 18 years old requiring RSI were eligible for enrolment. For the present retrospective analysis, we included consecutive children who received IV remifentanil bolus for a RSI procedure, with at least one of the following risk factors for pulmonary aspiration: bowel occlusion, recent vomiting or nausea, post traumatic ileus, severe chronic gastric dysfunction, insufficient preoperative fasting period (less than 6 hours). Exclusion criteria included the planned use of neuromuscular blocking agent for RSI, preoperative hypoxemia, hemodynamic shock, or difficult intubation risk factors.

### Usual per operative procedures

On arrival to the operating room, an IV catheter was inserted, if not already present. Baseline blood pressure, heart rate and pulse oximetry were routinely recorded. Remifentanil was injected as an intravenous bolus in combination with propofol as a hypnotic agent. The dose of each drug was predetermined by the anaesthesiologist in charge based on the patient’s body weight.

Intubation was performed by senior anaesthesiologists, residents with over a year of experience in anaesthesiology or nurses specialized in anaesthesiology, after oxygen preoxygenation in accordance with current guidelines. The choice to use prophylactic IV atropine (20µg/kg) during induction to prevent bradycardia was made by the anaesthesiologist in charge.

In the event of an early complication (within ten minutes of RSI), the use of rescue drugs or emergency ventilation techniques was notified.

### Collected Data

All data included in the study come from the “Vigilance Anaesthetic in the PeriOperative period” (VAPO) questionnaire. The VAPO is a prospective collection of perioperative anaesthetic data, integrated directly into the computerised patient chart. The VAPO has been used in our paediatric centre for several years for all anaesthesia procedures to document the perioperative anaesthetic management, monitor patients at risk of complications and track post-operative outcomes. A special section was added to obtain data on the use of IV bolus remifentanil in the RSI procedure.

The study was approved by an institutional review board in April 2022 (IRB Montpellier University Hospital – IRB-MTP_2022_04_202201075). No consent collection was required for this retrospective observational study.

Patient characteristics included: age, sex, weight, American Society of Anaesthesiologists classification (ASA). Surgery data included the timing of surgery (urgent or scheduled) and the type of surgery (urologic, digestive, orthopaedic, neurosurgery or other). Intubation data included: risk factors for pulmonary aspiration, induction drugs and their doses, use of prophylactic atropine during induction, use of emergency drugs after intubation. The use of direct or indirect laryngoscopy, Cormack Lehane grade, number of laryngoscopy attempts prior to intubation, vocal cord position during the intubation procedure, cough response during intubation and operator satisfaction (very satisfied, satisfied, moderately satisfied or unsatisfied) were also reported.

### Outcomes

The primary outcome was successful tracheal intubation at the first attempt without major complications following RSI with IV bolus remifentanil used in combination with a hypnotic agent. **A** successful intubation at the first attempt was defined as a successful endotracheal tube placement during the first laryngoscope insertion. Complications were collected up to ten minutes after the administration of induction drugs. Major complications were defined as: hypotension requiring more than one injection of ephedrine and IV crystalloid administration (20 ml/kg), bradycardia requiring more than one dose of IV atropine (20 µg/kg), hypoxemia below 90% requiring face mask ventilation prior to intubation or manual ventilation following intubation, pulmonary aspiration. Minor complications were defined as: an episode of hypotension reversible after just one injection of ephedrine or IV crystalloid fluid administration (20ml/kg), an episode of reversible bradycardia after a single dose of atropine or oxygen desaturation below 90%, quickly reversible after intubation and initiation of ventilation.

The secondary outcomes of the study were to evaluate the factors associated with intubation failure at the first attempt, the description of complications and the effect of preventive treatments.

### Statistical analysis

#### Primary outcome

The proportion of successful tracheal intubations at the first attempt without major complications was presented as percentage and its 95% Confidence Interval (CI).

#### Secondary outcomes

##### Failed intubation at first attempt

To evaluate factors associated with failed intubation at first attempt, a univariate analysis was performed on patient characteristics, the timing and the type of the surgery, remifentanil and propofol doses and use of prophylactic atropine. Count data, such as the number of laryngoscopy attempts, were compared using a Poisson regression. The comparison between groups (success *vs* failed at first attempt) on categorical data were performed using the χ2 test or the Fisher exact test, as appropriate. Continuous data were compared with the Wilcoxon test. Factors with a P-value under 0.20 or clinically relevant factors were integrated to a multivariate logistic regression using a backward procedure. Independent predictors of failed intubation were presented with adjusted odds ratios (OR) and its 95% confidence interval.

##### Complications and prophylactic treatment

After a descriptive analysis of complications, a post hoc analysis was performed to evaluate the benefit of prophylactic atropine using a propensity score matching methodology. Patients in the initial population were divided into two groups according to the administration or not of prophylactic atropine. A univariate analysis was conducted to compare baseline data between groups. Comparisons of categorical data among groups were performed using the χ2 test or the Fisher exact test, as appropriate. Continuous data were compared with a Wilcoxon test.

A multivariate logistic regression on the initial population was conducted on data selected in the univariate analysis, with a threshold of 0.2 to generate a propensity score. We used nearest neighbour matching without replacements and adopted a calliper equal to 0.1 to create matched pairs. The comparisons of outcomes between groups in the final population were evaluated using the Mc Nemar test for paired qualitative data.

Descriptive continuous variables were presented as means (SD) or medians (IQR) and categorical data as numbers (%).

P-values below 0.05 were considered statistically significant. The statistical analysis was carried out using SAS Enterprise version 8.3 (SAS Institute, Cary, NC).

## RESULTS

### Study population

We included 267 children at risk for pulmonary aspiration who underwent GA using a “remifentanil RSI” procedure.

The mean age was 7.8 +/- 4.4 years old. Age ranged from one day to sixteen years, including 21 (7.9%) under 1 year. Patient characteristics are presented in Table 1. The main risk factors for pulmonary aspiration were digestive occlusion, recent vomiting or post traumatic ileus, reported in 81.3% of cases (217 patients). Digestive or urologic surgery was the most frequent procedure, reported in 56.9% of cases (152 patients). The hypnotic agent used in combination with remifentanil was propofol in 100% of cases. The median (quartile) dose of bolus remifentanil was 3.1 (2.9-3.5) µg/kg. Direct laryngoscopy was used in 208 cases (78.2%).

**Table 1.** Patient characteristics.

| Characteristics | Remifentanil n=267 |
| --- | --- |
| Age, mean (SD), years | 7.84 (4.39) |
| < One year n (%) | 21 (7.9) |
| > One year n (%) | 246 (92.1) |
| Male n (%) | 172 (64.4) |
| Weight, mean (SD), kg | 30.04 (16.95) |
| American Society of Anaesthesiologists Physical Status Classification System score (ASA) n (%) |  |
| I | 179 (67) |
| II | 53 (19.9) |
| III | 33 (12.4) |
| IV | 2 (0.8) |
| Timing of surgery n (%) |  |
| Urgent or semi urgent | 249 (93.3) |
| Scheduled | 18 (6.7) |
| Type of surgery n (%) |  |
| Digestive, urologic | 152 (56.9) |
| Orthopaedic | 73 (27.3) |
| Others: Neurosurgery, endoscopic procedures | 42 (15.7) |
| Risk factors for pulmonary aspiration n (%) |  |
| Bowel occlusion, recent vomiting | 154 (57.7) |
| Post traumatic ileus | 63 (23.6) |
| Others: severe chronic gastric dysfunction, insufficient preoperative fasting period | 50 (18.3) |
| Remifentanil dose, mean (SD), µg/kg | 3.19 (0.66) |
| Propofol dose, mean (SD), mg/kg | 6.31 (1.88) |
| Prophylactic atropine n (%) | 114 (42.7) |
| Laryngoscopy n (%) (n=266) |  |
| Direct Laryngoscopy | 208 (78.2) |
| Video Laryngoscopy | 58 (21.8) |
| Cormack-Lehane grade n (%) (n=266) |  |
| 1: Full glottis view | 258 (97) |
| 2: Partial view of the glottis | 7 (2.6) |
| 3: Only epiglottis seen, no view of the glottis seen | 1 (0.4) |

### Primary outcome

Successful intubation at the first attempt without major complications was accomplished in 92.9% (95% CI, 88.9-95.6) of children (248/267 patients).

No major complication or pulmonary aspiration was observed during the study period.

### Secondary outcomes

Intubation failed at the first attempt in 19 children (7.1%). Successful intubation required two and three attempts in 16 (6%) and 3 (1.1%) patients, respectively. According to a significance level (p< 0.20) or clinically relevant factors in the univariate analysis (Table 2), 12 variables (Age younger than 1 year, weight, sex, ASA score, timing of surgery, type of surgery, risk factors for pulmonary aspiration, direct laryngoscopy, Cormack-Lehane grade, prophylactic atropine use, remifentanil dose and propofol dose) were used in the multivariate logistic regression. Only age less than one year was reported as an independent factor for failed intubation at first attempt (OR, 11.4; 95% CI, 3.7-35.1; p<0.001).

**Table 2:** Successful intubation at the first attempt in the univariate analysis.

| Characteristics | Successful intubation at the first attempt<br>n= 248 | Failure intubation at the first attempt n=19 | P value |
| --- | --- | --- | --- |
| Age, median (IQR), years | 8.00 (5.00 ; 12.00) | 1.00 (0.08 ; 8.00) | <0.01 |
| < One year n (%) | 13 (5.24) | 8 (42.11) | <0.01 |
| Weight, median (IQR), kg | 27.10 (18.80 ; 41.70) | 10.61 (3.83 ; 20.75) | <0.01 |
| Male n (%) | 157 (63.31) | 15 (78.95) | 0.17 |
| ASA score n (%) |  |  | <0.01 |
| I | 172 (69.35) | 7 (36.84) |  |
| II | 48 (19.35) | 5 (26.32) |  |
| III | 27 (10.89) | 6 (31.58) |  |
| IV | 1 (0.40) | 1 (5.26) |  |
| Timing of surgery n (%) |  |  | 0.13 |
| Scheduled | 15 (6.05) | 3 (15.79) |  |
| Urgent or semi urgent | 233 (93.95) | 16 (84.21) |  |
| Type of surgery n (%) |  |  | 0.04 |
| Orthopaedic surgery | 72 (29.03) | 1 (5.26) |  |
| Digestive, urologic surgery | 139 (56.05) | 13 (68.42) |  |
| Other surgeries | 37 (14.92) | 5 (26.32) |  |
| Risk factors for pulmonary aspiration n(%) |  |  |  |
| Bowel occlusion, recent vomiting | 140 (56.45) | 14 (73.68) | 0.14 |
| Post traumatic ileus | 62 (25.00) | 1 (5.26) | 0.05 |
| Other risk factors | 46 (18.55) | 4 (21.05) | 0.76 |
| Direct laryngoscopy n (%) | 194 (78.54) | 14 (73.68) | 0.57 |
| Cormack-Lehane grade n (%) |  |  | <0.01 |
| 1 | 243 (98.38) | 15 (78.95) |  |
| 2 | 4 (1.62) | 3 (15.79) |  |
| 3 | 0 (0.00) | 1 (5.26) |  |
| Prophylactic Atropine n (%) | 104 (41.94) | 10 (52.63) | 0.36 |
| Remifentanil dose, median (IQR),<br>µg /kg | 3.10 (2.90 ; 3.40) | 3.40 (2.80 ; 3.90) | 0.36 |
| < 3.1 µg /kg n (%) | 136 (54.84) | 12 (63.16) | 0.48 |
| Propofol dose, median (IQR), mg/kg | 6.00 (5.10 ; 7.50) | 6.00 (4.20 ; 7.00) | 0.33 |

Vocal cords were in abducted position in 245 (92.8%) children, and a coughing reaction when passing through the cords was noted in 67 (25.8%) children. Operators rated intubation conditions as very satisfactory in 210 (81.7%) children and satisfactory in of 39 (15.2%) children.

Forty-two (15.7%) children had at least one minor complication during the 10 minutes following induction. Thirty-eight (14.2%) patients presented an episode of minor hypotension, which was corrected by a single injection of ephedrine. Three episodes (1.1%) of bradycardia were recorded in older children: two were 13 years old and the third was 16 years old. Two episodes of bradycardia were associated with minor hypotension. Three patients (1.1%) presented an episode of desaturation below 90%. All were aged less than one year. For two children, the VAPO reported an episode of bronchospasm linked to selective intubation, quickly corrected by adjusting the tracheal tube position. The third episode of desaturation was linked to difficulties with tracheal catheterization, requiring a change of operator.

### Post hoc analysis

Prophylactic IV atropine was administered in 114 (42.7%) children. The number of children with minor hypotension was significantly lower in the group with prophylactic atropine than in the group without atropine (8.8% versus 18.3%; p=0.03).

A multivariate logistic regression on the initial population was conducted to generate a propensity score. The model included covariables associated with the administration of atropine with a *P* < 0.2 or a clinically relevant factor: age, weight, timing of surgery and type of surgery. Two hundred and twenty-four patients were used in the final population to analyse the impact of atropine in two comparable groups (Table 3).

**Table 3:**
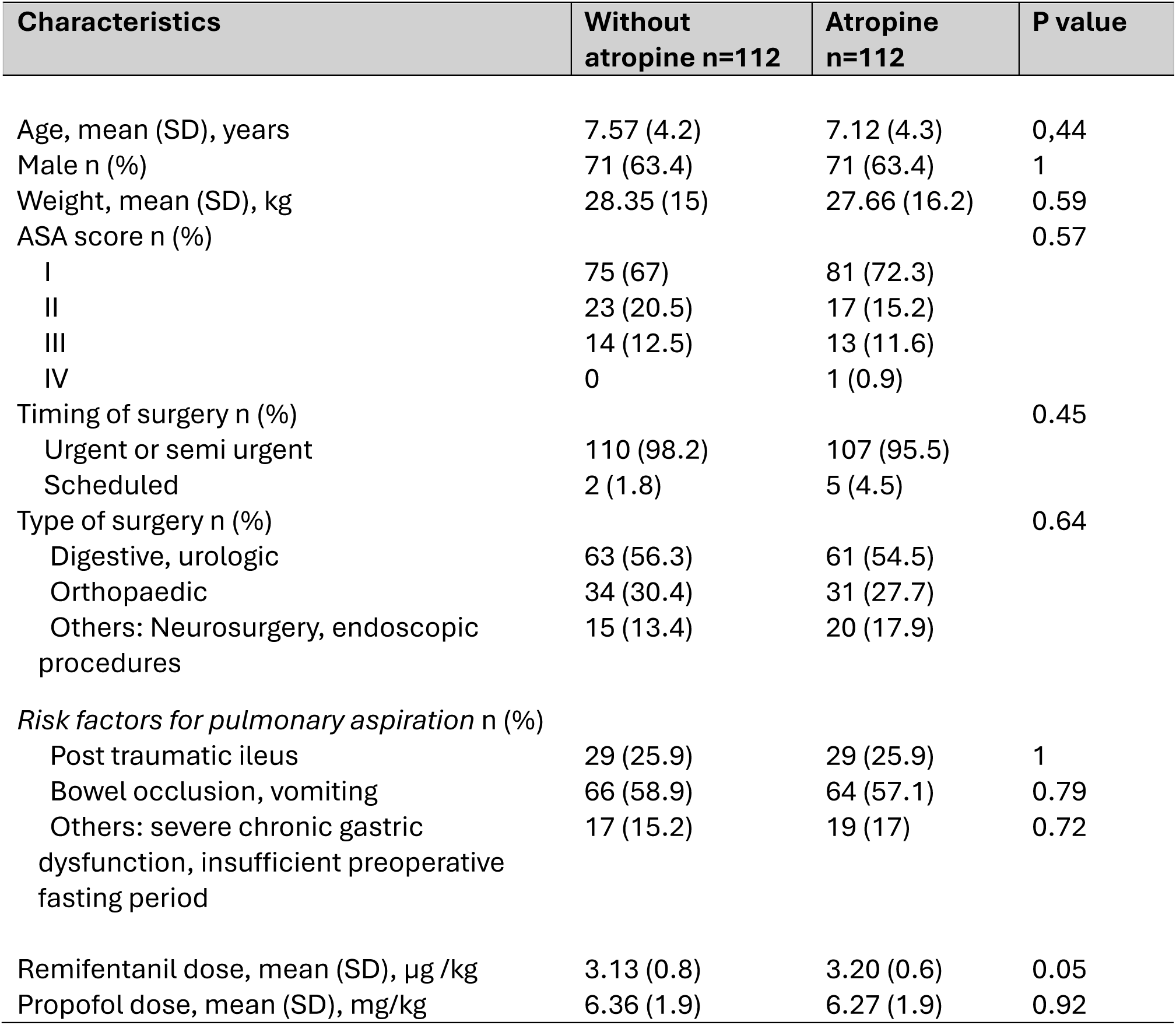
Patient characteristics after propensity score matching for prophylactic atropine.

| Characteristics | Without atropine n=112 | Atropine n=112 | P value |
| --- | --- | --- | --- |
| Age, mean (SD), years | 7.57 (4.2) | 7.12 (4.3) | 0,44 |
| Male n (%) | 71 (63.4) | 71 (63.4) | 1 |
| Weight, mean (SD), kg | 28.35 (15) | 27.66 (16.2) | 0.59 |
| ASA score n (%) |  |  | 0.57 |
| I | 75 (67) | 81 (72.3) |  |
| II | 23 (20.5) | 17 (15.2) |  |
| III | 14 (12.5) | 13 (11.6) |  |
| IV | 0 | 1 (0.9) |  |
| Timing of surgery n (%) |  |  | 0.45 |
| Urgent or semi urgent | 110 (98.2) | 107 (95.5) |  |
| Scheduled | 2 (1.8) | 5 (4.5) |  |
| Type of surgery n (%) |  |  | 0.64 |
| Digestive, urologic | 63 (56.3) | 61 (54.5) |  |
| Orthopaedic | 34 (30.4) | 31 (27.7) |  |
| Others: Neurosurgery, endoscopic procedures | 15 (13.4) | 20 (17.9) |  |
| *Risk factors for pulmonary aspiration n (%)* |  |  |  |
| Post traumatic ileus | 29 (25.9) | 29 (25.9) | 1 |
| Bowel occlusion, vomiting | 66 (58.9) | 64 (57.1) | 0.79 |
| Others: severe chronic gastric dysfunction, insufficient preoperative fasting period | 17 (15.2) | 19 (17) | 0.72 |
| Remifentanil dose, mean (SD), µg /kg | 3.13 (0.8) | 3.20 (0.6) | 0.05 |
| Propofol dose, mean (SD), mg/kg | 6.36 (1.9) | 6.27 (1.9) | 0.92 |

Minor complications were significantly lower in the group with prophylactic atropine than without (9.8% vs 17.9 %; p<0.001). The use of emergency drugs was significantly more frequent in the group without prophylactic atropine (17 % vs 8 %; p<0.001) (Table 4).

**Table 4:** Comparison of complication rates after propensity score matching for prophylactic atropine.

| **Characteristics n (%)** | **Without atropine**<br>**n=112** | **Atropine**<br>**n=112** | **<u>P value</u>** |
| --- | --- | --- | --- |
| Minor complication | 20 (17.9) | 11 (9.8) | **<0.001** |
| Episode of minor hypotension | 19 (20) | 10(8.9) | **<0.001** |
| Episode of minor bradycardia | 2 (1.8) | 0 | **<0.001** |
| Episode of saturation <90% | 0 | 2 (1.8) | **<0.001** |
| Emergency drugs<br>(ephedrine, atropine) | 19 (17) | 9 (8) | **<0.001** |

## DISCUSSION

This study is the first to report the use of remifentanil in combination with propofol for RSI in children. Our results showed a high success rate of 93.2% for tracheal intubation at the first attempt without major complications or pulmonary aspiration. In addition, a post hoc analysis revealed that the prophylactic use of atropine could be associated with a reduction in minor complications. This approach could be a valuable alternative in the anaesthetic management of paediatric patients at risk of aspiration.

The current study is the first to analyse the use of IV bolus remifentanil for RSI in a paediatric population. Previous studies (13–14), which reported excellent intubation conditions with remifentanil used in paediatrics, systematically excluded emergency procedures, often associated with a potential risk of aspiration. Blair et al. (13) showed that remifentanil at a dose of 3µg/kg provides similar intubation conditions to mivacurium. In the same way, Klemola et al. (14) showed that excellent intubation conditions were more frequent in the group receiving IV remifentanil at a dose of 4µg/kg than in the rocuronium group. In accordance, the average dose of bolus remifentanil used in the present study was 3.19 +/- 0.66 µg/kg. In addition, in the current study, the selected population was children at risk of aspiration, for whom practitioners were free to choose a classical or modified RSI protocol, including, or not, the planned use of a neuromuscular blocking agent. In the “remifentanil RSI” current cohort, we reported a successful tracheal intubation at the first attempt in 93% of patients. Our results are consistent with previous studies that reported intubation success rates after RSI between 78% (15) and 84% (16) with the use of neuromuscular blocking agents in paediatric populations. In another prospective cohort including 9297 children who underwent a classical RSI, Von Ungern-Sternberg et al. (17) found a success rate of 86% for intubation at the first attempt.

Regarding intubation conditions, while the vocal cords were in the abducted position in 92.8% of children, a coughing reaction during tube passage through the vocal cords was notified in 25.8% of cases. These results appear to be consistent with the study by Hannah et al (18), who reported no difference in the occurrence of a cough reaction between intubations with suxamethonium 1.2 mg/kg and remifentanil 4 µg/kg. Acceptable intubation conditions have been shown to increase with increasing doses of remifentanil (13;19). Considering the pharmacological properties of remifentanil, a low dose of remifentanil and a short a delay between the injection and the laryngoscopy could explain the coughing reaction observed.

In a recent adult study by Grillot et al. (20), remifentanil was compared to suxamethonium for RSI. The remifentanil group did not meet the criterion for non-inferiority regarding successful intubation without major complications (66% vs 71% in the remifentanil group vs the neuromuscular blocking agent group, respectively). RSI failure was attributed to the inability to intubate on the first attempt, while episodes of major hemodynamic instability, prolonged arrhythmia >30 s or cardiac arrest were reported in 20% of patients similarly in remifentanil and neuromuscular blocking agent groups. We chose to apply a similar primary outcome in the present study. In contrast with the adult study, we demonstrated a higher rate (93%) of successful intubation at the first attempt and no major complication or pulmonary aspiration in our paediatric cohort. As previously indicated, the paediatric population is significantly distinct from the adult population. In the present study, episodes of mild and transient hypotension were the most frequent minor complications, observed in 13.9% of cases. Such episodes of hypotension were reversible with a single injection of ephedrine or crystalloid fluid administration. In addition, hypotensive events were significantly less pronounced in the group that received prophylactic atropine, after propensity score matching. In accordance, Blair et al. (13) and Klemola et al. (14), who systematically used prophylactic IV atropine (at doses of 10 µg /kg or 15µg/kg) when using remifentanil for induction, did not report hypotension or bradycardia requiring treatment. It should be noted, however, that there is currently no consensus on what constitutes hypotension in the literature on paediatric anaesthesia. Wani et al. (21) studied variations in blood pressure in children aged 2 to 8 years in the 12 minutes following the induction of anaesthesia to investigate the incidence of post-induction hypotension. The incidence varied considerably based on the definition used, being 27.5% if the 5th percentile of the systolic blood pressure for age was used versus 57.2% (107 of 189 patients) when a 20% decrease of the systolic blood pressure from baseline was used. In adult classical RSI, a high rate of cardiorespiratory complications was reported, with 18% experiencing hypotension after GA induction (22).

Finally, the only independent factor for intubation failure at the first attempt was an age of less than 1 year (OR, 11.4; 95% CI, 3.7-35.1). This result reinforces the literature on paediatric airway management, which indicates that small children, particularly those aged under one year or weighing less than 10 kg, are at greatest risk (23).

Therefore, we suggest that the use of remifentanil could be considered as an alternative in a selected population of children at risk of pulmonary aspiration, provided it is administered, as in the present study, by a paediatric anaesthesia team trained in its use. Prophylactic IV atropine should also be considered.

The concept of RSI was introduced in 1951 to enhance the safety of patients at risk of pulmonary aspiration. The initial description remained unchanged for decades. The classical approach to RSI includes the use of rapid-onset neuromuscular blocking agents, and it is currently recommended in the latest paediatric guidelines (4). However, it appears to be associated with some complications. Recently, Engelhardt believed that RSI could present specific risks if applied without modification in paediatric anaesthesia (24). Controlled or modified approach to RSI may include gentle ventilation (7–10), with the aim in reducing the incidence of hypoxemia more than the incidence of pulmonary aspiration (22). It may also include other neuromuscular blocking agents or the administration of opioids (25). Indeed, several surveys over the past decades have evaluated RSI practices and revealed a wide variation in clinical practice, particularly in paediatric anaesthesia. (25–27). This evolution could be partially attributed to the potential adverse effects of muscular blocking agents in children, including prolonged paralysis, metabolic disturbance, post-operative respiratory complications and anaphylaxis. The national survey, conducted by Petitpain et al (28), revealed a significantly increased risk (19 to 22 times higher) of anaphylaxis with suxamethonium compared to other agents. Cases of severe anaphylaxis in children have been documented, emphasizing the need for caution and available alternative approaches in paediatric anaesthesia. The use of remifentanil for paediatric RSI provides an alternative in this context.

Limitations of our study include the absence of a control group and the analysis of retrospective data. However, the aim of the study was not to compare or substitute neuromuscular blocking agents, but rather to describe a new approach to RSI with remifentanil in terms of safety. Similarly, the outcomes of hypotension and bradycardia were subjective and depended on the decision of treatment by the anaesthesiologist, with no predefined threshold. However, as previously indicated, the definition of thresholds is a recurrent debate in paediatrics (21).

## CONCLUSION

This study presents new arguments on the feasibility and safety of using bolus IV remifentanil for RSI in a heterogeneous sample ranging from new-borns to adolescents over a two-year period. The use of IV bolus remifentanil could be considered a promising proof-of-concept for the development of future international guidelines for modified or controlled RSI in paediatric anaesthesia. A larger study and controlled trials in a selected population of children undergoing modified RSI should be carried out.

## Data Availability

All data produced in the present work are contained in the manuscript

## Notes

### Competing Interest Statement

The authors have declared no competing interest.

### Funding Statement

This study did not receive any funding

### Author Declarations

The study was approved by an institutional review board in April 2022 (IRB Montpellier University Hospital — IRB-MTP_2022_04_202201075).

